# Potential Predictive Factors for Breast Cancer Subtypes from a North Cyprus Cohort Analysis

**DOI:** 10.1101/19010181

**Authors:** Ayse Ulgen, Özlem Gürkut, Wentian Li

## Abstract

**Purpose:** We present a first epidemiological survey from North Cyprus to determine predictive factors for breast cancer subtypes.

**Methods:** More than 300 breast cancer patients, 90% of them having subtype information, are surveyed from the State Hospital in Nicosia between 2006 – 2015 for their demographic, reproductive, genetic, epidemiological factors. The breast cancer subtypes, Estrogen receptor (ER) +/-, Progesterone receptor (PR) +/-, and human epidermal growth factor 2 (HER2) +/- status, are determined. Single and multiple variable, regularized regressions, with predictive factors as independent variables, breast cancer subtypes as dependent variables are conducted.

**Results:** Our cohort differs significantly from larger cohorts (e.g., Breast Cancer Family Registry), in age, menopause status, age of menarche, parity, education, oral contraceptive use, breastfeeding, but the distribution of breast subtypes is not significantly different. Subtype distribution in our cohort is also not different from another Turkish cohort. We show that the ER+ subtype is positively related to age/post-menopause; ER+/PR+ is positively associated with age, but negatively associated with cancer stage; HER2+, which is negatively correlated with ER+ and ER+/PR+, is positively related to cancer stage but negatively associated with age/post-menopause.

**Conclusion:** Assuming ER+ and ER+/PR+ to have better prognostic, HER+ to have worse prognostic, then older age and postmenopause seem to be beneficial, smoking and family history of cancer seem to be detrimental. Next steps include looking at potential biomarkers and using cure models to determine long-term survivors.

## Introduction

Breast cancer is the most common type of cancer diagnosed in the Western part of the world. In Europe, more than 523,000 women were diagnosed with breast cancer in 2018 and more than 138,000 women died from it [1]. World-wide, close to 2 million women are diagnosed with breast cancer each year and approximately 30% die from this disease. Breast cancer is largely viewed as a disease predominantly influenced by risk factors related to lifestyle [2] though through the twin studies of heritability of breast cancer, genetic contribution can still be significant [3]. Recent work to combine contribution from many genetic variants to breast cancer achieves an above 60% area-under-receive-operator-curve prediction rate [4, 5], and 20% variance explained [6].

Female hormones may affect breast cancer, and their status have been used to classify breast cancers. In particular, estrogen receptor positive (ER+) or negative (ER-), progesterone receptor positive (PR+) or negative (PR-), human epidermal growth factor 2 positive (HER2+) or negative (HER-) are the major classification schemes of breast cancer subtypes. It has been shown that ER+/-, PR+/-, HER+/- breast cancer subtypes have different clinical features [7], the cancer etiology of these subtypes can be heterogeneous, and treatment strategies also diverge. In particular, hormone receptor-positive (ER+ or PR+) subtype should expect good prognosis, using drugs like Tamoxifen/Nolvadex Similarly, the more aggressive HER2+ subtype can be treated successfully with drugs like Trastuzumab/Herceptin. In contrast, the triple-negative subtype (ER-PR-HER2-) poses challenges in treatment strategies [8].

Although international and national studies of breast cancer have been conducted with large sample sizes, such as BCFR (*www.bcfamilyregistry.org*), GICR (*gicr*.*iarc*.*fr*), BCSC (*www.bcsc-research.org*), there has never been a breast cancer survey on the subtype distributions, potentially explanatory variables, and correlation between these variables and breast cancer subtypes, in North Cyprus (though there are some studies in Turkey [9]). To fill this gap, we present a first epidemiological survey of around 300 breast cancer patients from North Cyprus – among them around 230 Turkish Cypriots.

We collected and analyzed reproductive (age of menarche, number of children (zero for nulliparity), menopause status, hormone therapy, oral contraceptive use, breastfeeding, left/right breast with cancer), demographic (age at diagnosis, education level, housewife/employed), genetic (first relative having cancer), and epidemiological (smoking, other cancers) information. Most of these factors are known to be risk factors for breast cancer, e.g., early menarche, late menopause, nulliparity, long hormone replacement therapy, older age, family history of breast cancer, but it is unclear which factor is predictive for breast cancer subtypes.

Our analysis strategy is as follows: We treat ER, PR, HER2 as dependent variables, and other as independent variables. As we do not have control (noncancer) samples, it is a case-only analysis or subtypes-with-case analysis [10, 11]. The first analysis is to compare our independent and dependent variables distribution with a major public breast cancer databases. Second, correlation between the cancer subtypes are determined. Third, uni-variate, multiple, and regularization logistic regression are performed to detect any factor-subtype association, i.e., to identify potential predictive factors for breast cancer subtypes. We will show that though there are some minor surprises, our cohort conforms with some other studies concerning predictive factors of breast cancer subtypes.

## Methods

### Sample collection

We included 324 samples (321 female, 3 male) collected retrospectively from the Dr. Burhan Nalbantoğlu State Hospital (BNSH) in Nicosia, North Cyprus between 2006–2015, mostly from years 2011–2015 (93%). This represented around 40% of total beast cancer cases that exist in the archives during this period. The data consists of reproductive factors, histology and biomarker information such as the ER, PR, and HER2 status. Permission was obtained from the Ministry of Health from the Turkish Republic of Cyprus for scientific use of the data. Additionally, ethical approval from the Eastern Mediterranean University Ethics Committee in Famagusta was granted to conduct the study. IRB approval number was AAAP8950. Patient consent forms were not required. Telephone interviews were made when necessary to collect information from patients to fill in the missing factor values.

For these cases, pathologists from the BNSH ascertained ER and PR status from patient tumor tissue using immunohistochemistry (IHC) and/or pathology reports using a standardized protocol and pathology reporting forms. For all cases, HER2 status available (around 290) was provided from patient medical reports. Where tumor tissue was available, pathologists used IHC testing for ER and PR, and categorized tumors as ER and PR positive if ≥ 10% of tumor cells stained positive. When the ER or PR +/- status is not labeled, but with a specific percentage, we treat it as unknown. Menopausal and other information were extracted either from the medical records (with guidance/approval from an oncologist) or by telephone interviews.

### Pre-processing of data

We remove the three male samples, reducing the sample size from 324 to 321. For hormone receptor status, if the left/right breast has different value, it is labeled as NA (unknown). Also, if the hormone receptor status is not binarized but represented by a percentage, it is labeled as NA.

Other re-coding of the data include: (smoking) seldom=0, quit=1, x-number-pocket=1; (family history) first degree relatives are parents, children, and siblings; (other cancer) anything not “no” is considered as yes (including metastasis); (education) 0,1,2,3 are for no school, primary/middle school, high school, college or more; (housewife/employment) retied is considered as the same as employed; (tumor stage) “high stage” is considered as 3, inoperable is considered as 4, A/B/C are ignored; (invasive cancer) IDC(invasive ductal carcinoma)/ICC(invasive cribriform cancer)/ISC(invasive secretory cancer) are invasive, everything else is not invasive. Of the 321 samples, 300 have ER or PR information, 291 have HER2 information; all have age information, but only 214, 215, 217, 222 patients have breastfeeding, age of menarche, use of oral contraceptive, use of hormone therapy information. The amount of missing data for other factors is given in Tables 1,2. An independent variable is removed if the missing rate is too high (e.g., > 0.2). The rest of missing value is imputed from the known variable value (e.g., if x is the independent variable, two values are missing, they are replaced by (R code): *sample(x[!is*.*na(x)][1:2])*.

**th Table 1.** Factors that are distributed differently between North Cyprus cohort and BCFR. Factors that are significantly different between North Cyprus cohort and BCFR: age at diagnosis, post or pre menopause status, age at menarche (first occurrence of menstruation), parity (number of births), education level (HS: high school), oral contraceptive use, beast feeding. pv(++ vs - -) is the Fisher’s test p-value comparing the North Cyprus ER+PR+ vs ER-PR-group. pv (NC vs BCFR) is the Fisher’s test p-value comparing North Cyprus and BCFR group. Missing data (NA) are not counted in calculating percentage and not used in Fisher’s test. All p-values smaller than 0.001 (this threshold is recommended in [37]) are marked by boldface.

| factor | North Cyprus |  |  |  | BC Fam Registry |  | NC vs BCFR pv |  |  |
| --- | --- | --- | --- | --- | --- | --- | --- | --- | --- |
|  | whole<br>n=321 | ER+PR+<br>n=204 | ER-PR-<br>n=64 | ++vs- -<br>pv | ER+PR+<br>n=2486 | ER-PR-<br>n=920 | all | ++ | - - |
| age | 57.4 ± 12.8 | 58.7 ± 12.5 | 55.3 ± 13 | 0.06 | 47.1 ± 9.3 | 44.5 ± 9.8 | | **6E-29** | **1E-8** |
| menopause | 201 (64.8%) | 136 (68.3%) | 36 (57.1%) | 0.13 | 951(40%) | 310(35%) | **5E-18** | **1E-14** | **6E-4** |
| not | 109 (35.2%) | 63 (31.7%) | 27 (42/9%) |  | 1431 (60%) | 574 (65%) |  |  |  |
| NA | 11 | 5 | 1 |  |  |  |  |  |  |
| menarche | 13.14 ± 1.31 | 13.11 ± 1.30 | 13.32 ± 1.25 | 0.3 | | | | | |
| age≤ 11 | 19 (8.8%) | 12 (8.5%) | 3 (5.7%) | 0.43 | 528(22%) | 183(21%) | **1E-5** | **3E-4** | 6E-3 |
| 12 | 54 (25.1%) | 40 (28.4%) | 11 (20.8%) |  | 590(24%) | 215(24%) |  |  |  |
| ≥ 13 | 142 (66%) | 89 (63.1%) | 39 (73.6%) | | 1317(54%) | 482(55%) | | | |
| NA | 106 | 63 | 11 |  |  |  |  |  |  |
| parity | 2.34 ± 1.42 | 2.47 ± 1.5 | 2.17 ± 1.18 | 0.1 | | | | | |
| no.kid=0 | 31 (10%) | 21 (10.5%) | 6 (9.4%) | 0.44 | 565 (23%) | 191(21%) | **9E-8** | **8E-5** | 0.04 |
| 1-2 | 159 (51.1%) | 91 (45.5%) | 35 (54.7%) |  | 1015(41%) | 391(42%) |  |  |  |
| ≥ 3 | 121 (38.9%) | 88 (44%) | 23 (35.9%) | | 906(36%) | 338(37%) | | | |
| NA | 10 | 4 | 0 |  |  |  |  |  |  |
| edu: < HS | 174(56.5%) | 113 (57.1%) | 33 (51.6%) | 0.47 | 710(29%) | 289(32%) | **1E-20** | **4E-15** | **0.002** |
| ≥ HS | 134(43.5%) | 85 (42.9%) | 31 (48.4%) | | 1740(71%) | 602(68%) | | | |
| NA | 13 | 6 | 0 |  |  |  |  |  |  |
| OC use | 76(35%) | 46 (32.4%) | 19(35.8%) | 0.73 | 1795 (73%) | 680 (77%) | **3E-31** | **6E-23** | **5E-10** |
| no | 141(65%) | 96 (67.6%) | 34(64.2%) |  | 648 (27%) | 198 (23%) |  |  |  |
| NA | 104 | 62 | 11 |  |  |  |  |  |  |
| breastfeed | 168(78.5%) | 112(80.6%) | 38 (71.7%) | 0.24 | 1359 (55%) | 454 (50%) | **2E-13** | **1E-9** | 0.003 |
| no | 46 (21.5%) | 27 (19.4%) | 15 (28.3%) |  | 1105 (45%) | 448(50%) |  |  |  |
| NA | 107 | 65 | 11 |  |  |  |  |  |  |

**Table 2.** Other factors. Similar to Table 1, a list of factors that are not significantly different between the North Cyprus cohort and BCFR: hormone therapy (HT), and family breast cancer history. Other factors don’t have available data in BCFR: tumor stage, whether the patient has other cancers, smoking, left (L) or right (R) breast or both with cancer, housewife or employed, invasive cancer. (note 1) never or former menopausal hormone therapy use.

| factor | North Cyprus |  |  |  | BC Fam Registry |  | NC vs BCFR pv |  |  |
| --- | --- | --- | --- | --- | --- | --- | --- | --- | --- |
|  | whole<br>n=321 | ER+PR+<br>n=204 | ER-PR-<br>n=64 | ++ vs - -<br>pv | ER+PR+<br>n=2486 | ER-PR-<br>n=920 | all | ++ | - - |
| HT | 24 (19.5%) | 14 (9.7%) | 7 (13%) | 0.6 | 424 (18%) | 111 (13%) | 0.03 | **0.0093** | 1 |
| no | 198 (80.5%) | 131 (90.3%) | 47 (87%) |  | 1955<sup>1</sup> (82%) | 758<sup>1</sup> (87%) |  |  |  |
| NA | 99 | 59 | 10 |  |  |  |  |  |  |
| fam-hist | 85 (26.5%) | 55 (27%) | 19 (29.7%) | 0.75 | 714 (9%) | 244 (27%) | 0.65 | 0.63 | 0.56 |
| no | 236 (73.5%) | 149 (73%) | 45 (70.3%) |  | 1761 (29%) | 673 (73%) |  |  |  |
| stage : 1 | 60 (23.4%) | 41 (24.4%) | 10 (18.2%) | 0.55 |  |  |  |  |  |
| 2 | 135 (52.7%) | 89 (53%) | 30 (54.5%) |  |  |  |  |  |  |
| 3 | 51 (19.9%) | 31 (18.5%) | 14 (25.5%) |  |  |  |  |  |  |
| 4 | 10 (3.9%) | 7 (4.2%) | 1 (1.8%) |  |  |  |  |  |  |
| NA | 65 | 36 | 9 |  |  |  |  |  |  |
| otherC | 17 (5.3%) | 11 (5.4%) | 4 (6.3%) | 0.76 |  |  |  |  |  |
| no | 304(94.7%) | 193 (94.6%) | 60 (93.8%) |  |  |  |  |  |  |
| smoke | 84 (26.2%) | 53 (26%) | 20 (31.2%) | 0.42 |  |  |  |  |  |
| no | 237 (73.8%) | 151 (74%) | 44 (68.8%) |  |  |  |  |  |  |
| breast:L | 162 (50.6%) | 107 (52.5%) | 31 (48.4%) | 0.27 |  |  |  |  |  |
| R | 149 (46.6%) | 90 (44.1%) | 33 (51.6%) |  |  |  |  |  |  |
| both | 9(2.8%) | 7 (3.4%) | 0 (0%) |  |  |  |  |  |  |
| NA | 1 | 0 | 0 |  |  |  |  |  |  |
| housewife | 118 (42.8%) | 83 (45.9%) | 21 (36.8%) | 0.28 |  |  |  |  |  |
| employed | 158 (57.2%) | 98 (54.1%) | 36 (63.2%) |  |  |  |  |  |  |
| NA | 45 | 33 | 7 |  |  |  |  |  |  |
| invasive | 237 (75.5%) | 155(76.4%) | 48 (75%) | 0.87 |  |  |  |  |  |
| no | 77 (24.5%) | 48 (23.6%) | 16(25%) |  |  |  |  |  |  |
| NA | 7 | 1 | 0 |  |  |  |  |  |  |

### Turkish Cypriots and non-Turkish-Cypriots patients

Of the 321 samples, 314 of them report the birth country, where the majority are born in Cyprus (n = 233), 53 born in Turkey, and remaining 28 born in other countries, including UK, Turkmenistan, and Bulgaria. Although our analysis is not on genetic or ethnicity’s contribution to the breast cancer subtypes, and foreign-born does not automatically imply non-Turkish-Cypriots, we run all analyses twice: one on n=321 samples and another on n=233 Turkish-Cypriots only samples.

### Statistical programs used

All statistical analysis either used R 3.5.1 (www.r-project.org, released July 2018) or SPSS 17.0 (released 2008, Chicago: SPSS Inv.). The *Rtsne* R package is used for the t-SNE analysis (*github*.*com/jkrijthe/Rtsne*), with the default parameter setting (e.g., perplexity=30, dims=2). The *glmnet* R package (*web*.*stanford*.*edu/ hastie/glmnet*) [12] is used for the following regularized regression: least absolute shrinkage and selection operator, or LASSO (alpha=1, family=”binomial”), elastic net (alpha=0.5), ridge (alpha=1). The logistic regression is carried out by the standard R function: *glm(*… *family=binomial(link=”logit”))*, and the Fisher’s test was conducted using the standard R function *fisher*.*test*. The independent two-sample t-test between age distribution, one from raw data and another from summary statistics, was conducted using our customized R script.

## Results

### Visual inspection of the data by t-SNE

The t-distributed stochastic neighbor embedding (t-SNE) [13] is a popular method to represent high-dimensional data in 2 or 3 dimensions. We previously use the technique in other applications in biology/genomics [14, 15].

We use 3 dependent variables (ER, PR, HER2), 5 quantitative independent variables (age of diagnosis, age of menarche, number of children, education level, cancer stage), and 10 binary independent variables (left/right breast, menopause, first relative with cancer, other cancer, smoker, hormone therapy, oral contraceptive use, breastfeeding, housewife/employment, cancer invasiveness). The quantitative variables are standardized to have zero-mean and unit-variance (*z*-transformation).

Due to high missing rates for age of menarche (missing rate = 33%), hormone therapy (31%), oral contraceptive use (32%), breastfeeding (33%), we only keep samples who have information on these factors. This reduced the sample size from 321 to 211 for t-SNE plot. For these 211 patients, other missing data (of much lower missing rate) are imputed.

Fig.1 shows one run of t-SNE. Because ER, PR, HER2 are part of the input, it is not surprising that their values are well partitioned in the plot (e.g., ER+ and ER-samples). It can be seen that ER+ samples tend to PR+, and HER2-, ER-samples tend to be PR- and HER2+. The 7 samples with other cancers (including metastasis) form a distinct cluster from the rest of the samples. While ER, PR, HER2 values separate in up-down direction in Fig.1, other factors, such as menopause status, breastfeeding, age, etc. seem to be separated in (not completely) orthogonal direction.

**Figure 1:**
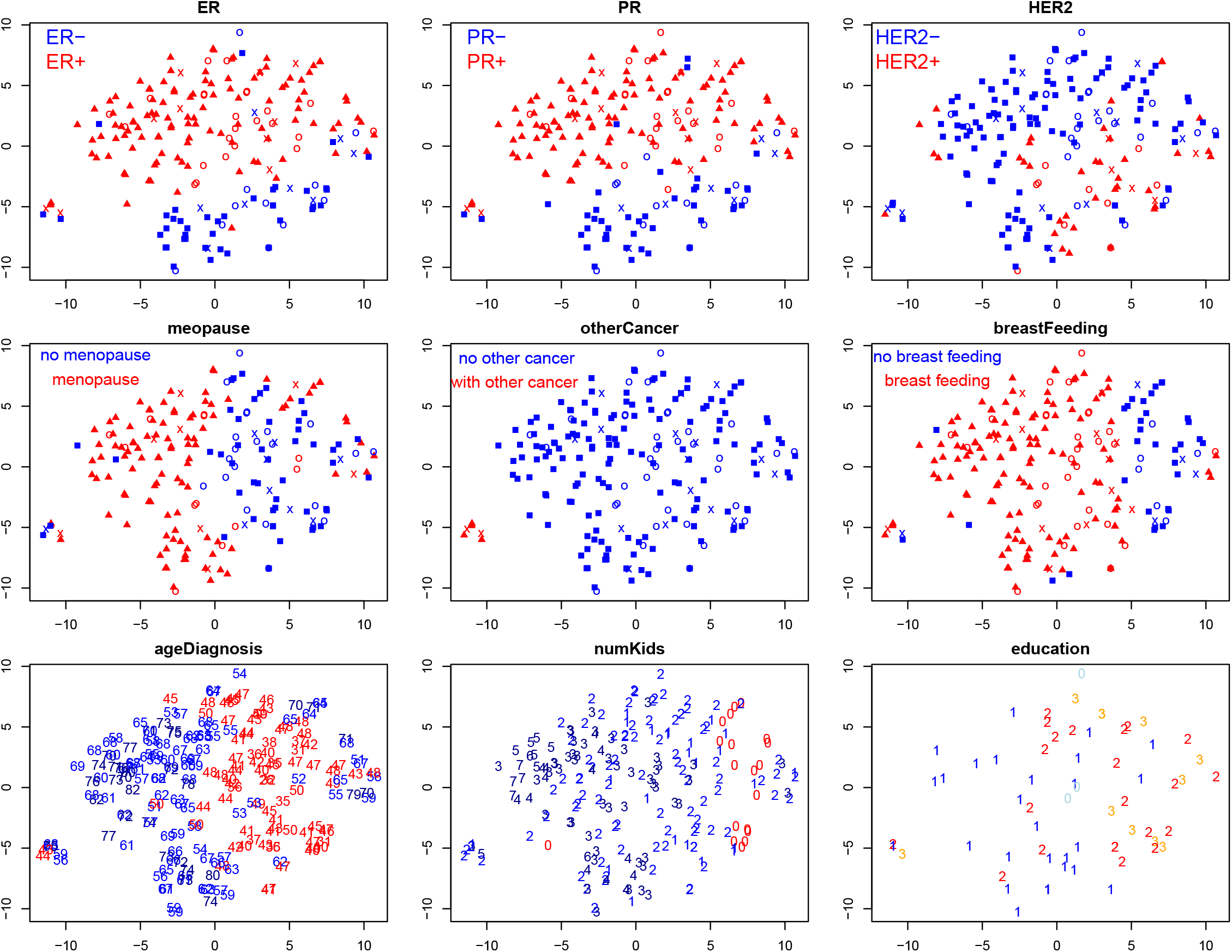
t-SNE plot of North Cyprus cohort samples. t-SNE plot of 211 breast cancer patients (out of 321 total) with enough non-missing factor values. The nine subplots are the same plot labeled with different information: ER subtype (red for ER+, blue for ER-), PR subtype, HER2 subtype, menopause status (post-menopause in red, pre-menopause in blue), if the patient has other cancer (red for yes, blue for no), breastfeeding (red for yes, blue for no), age of diagnosis (red if younger or equal of 50 years ago), parity/number of children, education level (0 for none, 1 for primary or middle school, 2 for high school, 3 for college or higher). The Turkey-born samples are marked with circle, and other foreign born samples are in crosses.

The non-Turkish-Cypriots are marked with different symbols (Turkey-born patients in circles, other foreign-country-born patients in crosses) in the top two rows of Fig.1. There is no evidence that the location of these points in the plot, or their collective features, are very different from the rest Turkish-Cypriots samples.

### Distribution of patient’s factors

Table 1 shows that our cohort (n=321) is distinct from the Breast Cancer Family Registry (BCFR) samples, which are mostly of USA/Canada/Australia origin, in several demographic or reproductive factors.

The North Cyprus cohort was older in age, had lower education level and less use of oral contraceptives, and had greater number of postmenopause subjects, lesser number of subjects with young (≤ 11) age at menarche, fewer nulliparous subjects, and greater number of breast-feeding subjects. The same analysis is carried out for n=233 Turkish-Cypriots patients only, and the conclusions remain the same (data not shown).

There are two explanations for these significant differences. The first is due to cultural and customary differences between countries (e.g., use of oral contraceptives). The second explanation is that our samples are collected from the state hospital, and a higher percentage of affluent patients may select to be treated at private hospitals, or hospitals overseas. The differences remain even for ER+/PR+ subgroup, and for ER-/PR-subgroup (though less significant due to smaller sample sizes).

Within our North Cyprus cohort, when the ER+/PR+ and ER-/PR-groups are compared on these factors, only ER-/PR- is slightly younger (t-test p-value = 0.06) (Table 1). Differences in other factors are not yet significant, either truly not different, or not enough sample size to prove the difference.

There are also some other factors distributed not very differently between our cohort and BCFR, as summarized in Table 2. These include hormone therapy usage and having a first-degree relative with cancer. The rest of factors in Table 2 do not have the corresponding information in BCFR, including other cancers, smoking status, left/right breast with cancer, housewife/employed, and cancer invasiveness. Only for ER+/PR+ subtype, the North Cyprus cohort is significantly less likely to have hormone therapy than the BCFR samples.

We also examine correlation between factors. Using all breast cancer patients without considering the subtypes, these correlations are observed: (1) patients who breast feed are less likely to go through hormone therapy (OR=7.1, Fisher p-value 9 ×10^−5^); (2) patients who work are more likely to smoke than housewives (OR=3.1, p-value= 1.3 ×10^−4^); (3) patients who work are more likely to be pre-menopause than housewives (OR=2.8, p-value= 1.3 ×10^−4^).

### Conversion of age to age-group

Age is a special factor different from others because it is a continuous variable spanning a wide range of values. Discretizing or categorizing a continuous variable is an involved topic by itself. Age is a well known target for such a categorization [16]. We partition samples into younger and older age-group by an age-threshold. When an age threshold is chosen, 2-by-2 count table can be constructed by the binary age group and binary breast cancer subtype. Age group versus breast cancer subtype association can be judged by Fisher’s test.

Fig.2 shows (top) (− log) Fisher’s p-value and (bottom) adds-ratio as a function of the age threshold for converting age to age group. For HER2, the best p-value is 0.0002 when age threshold is 42, and there is a broad range of age threshold (41-47) where the Fisher’s test is significant at 0.01 level. For ER, PR, ER/PR, this age threshold range also lead to some significant test results, indicating that patients younger than mid-40s may form a distinct group, that tend to ER, PR, ER/PR negative and HER2 positive. At age threshold 67-68, there is a second peak, indicating that patients older than that age tend to be ER, PR, ER/PR positive.

**Figure 2:**
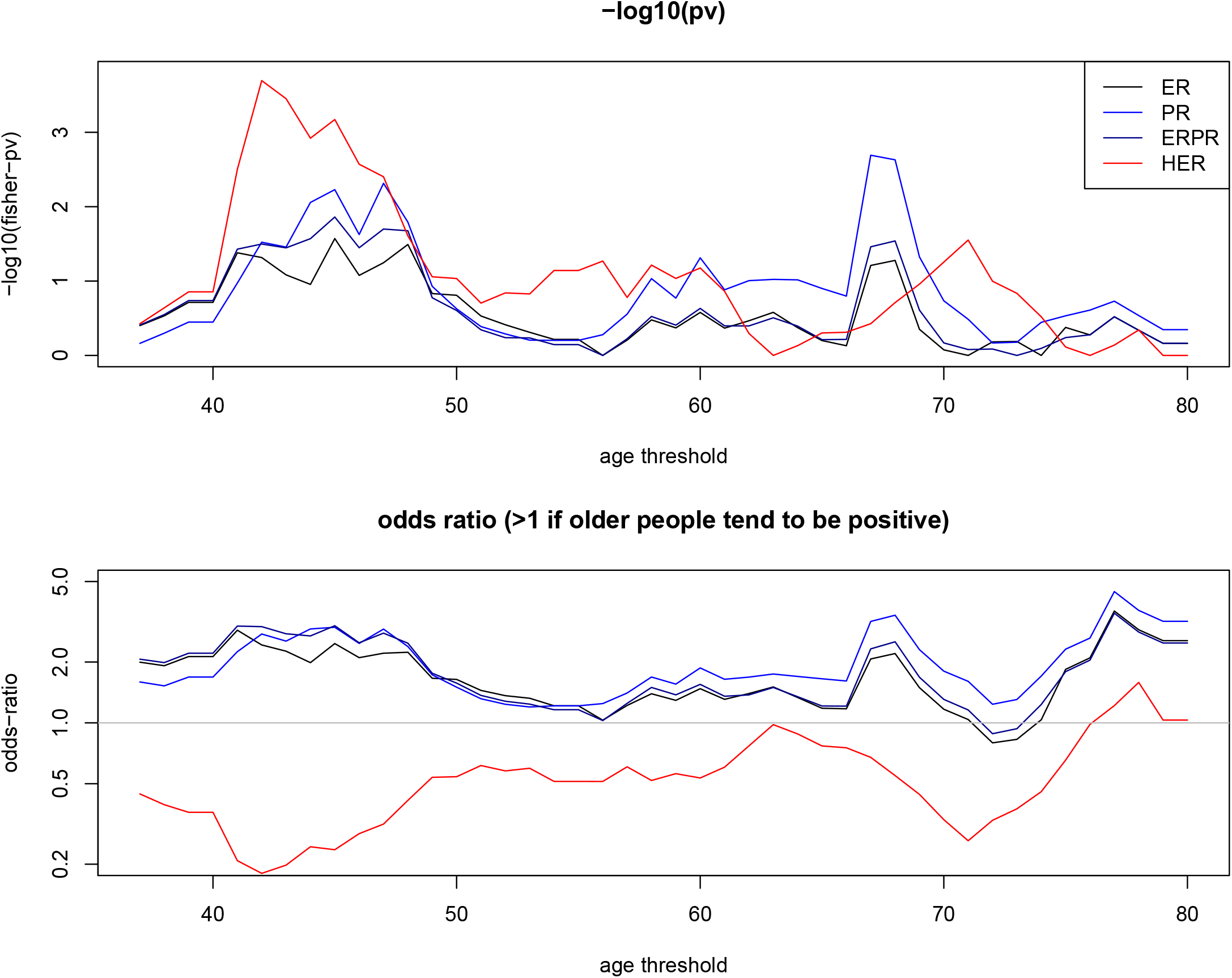
Tuning age threshold to convert age to age group. (top) Fisher’s p-value (in minus log with base 10) of age-group vs. breast cancer subtype (ER, or PR, or ER/PR, or HER2) as a function of the age threshold. (bottom) adds-ratio as a function of age threshold.

### Distribution of breast cancer subtypes

We compare the hormone receptor defined subtype distribution between the North Cyprus cohort and BCFR in Table 3. The count in each of the 8 ER/PR/HER2 subtypes in n=321 set and (Cyprus-born only) n=233 set is listed in Table 3(A)). These counts are not available for BCFR cohort [17], but the distribution by ER/PR subtypes, and by HER2/(ER-PR- and not) subtypes are available which is reproduced in Table 3(B) and (C).

**Table 3.**
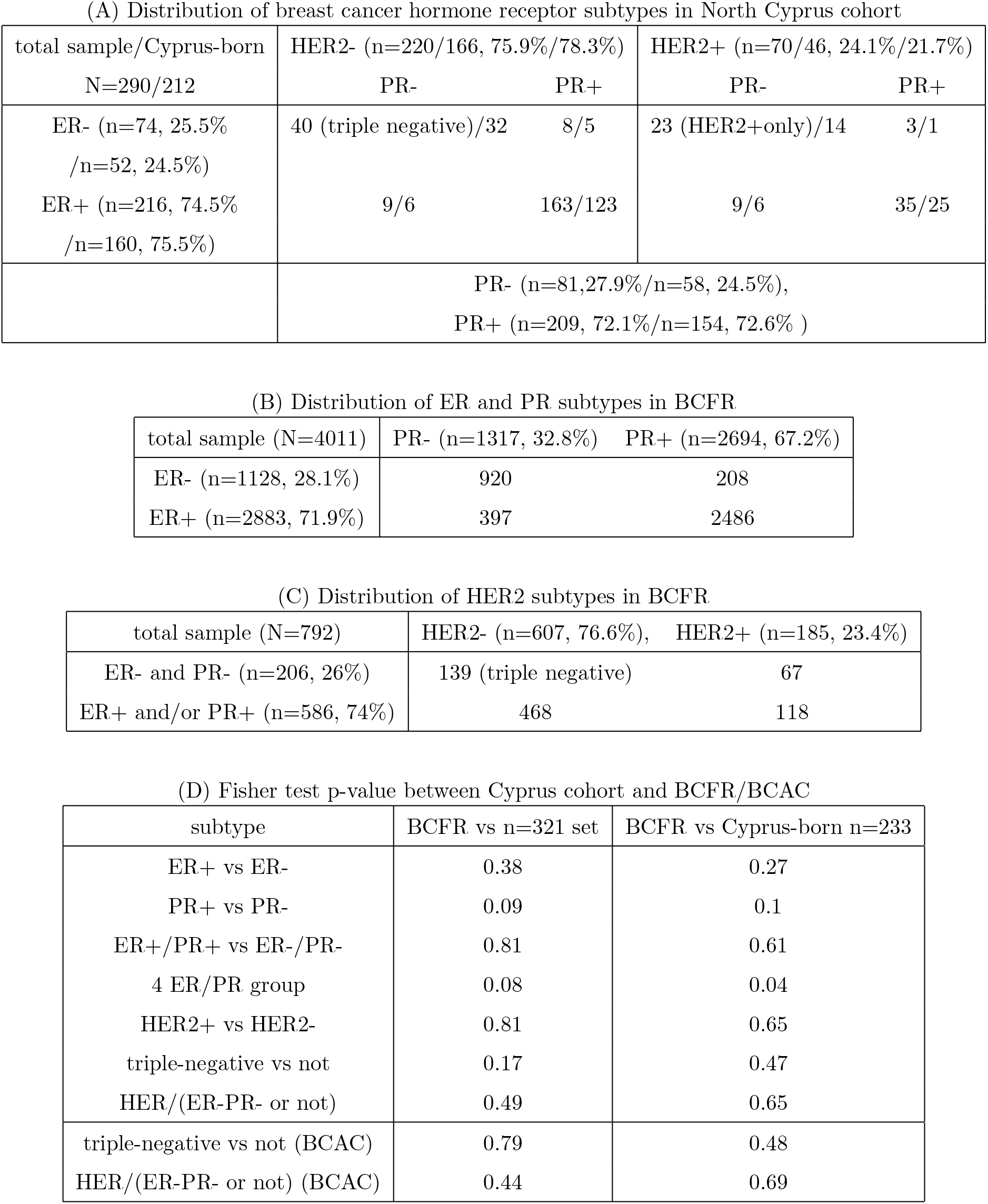
Count and testing of ER, PR, HER2 distributions. (A) Breast cancer subtype counts in the North Cyprus cohort (either using all samples or using Cyprus-born only samples). Hormone receptor positive (including luminal A and luminal B) consists of 8+9+163=180 counts (62.1%) if all samples are used; and (5+6+123)/212=63.2% if Cyprus-born samples are used. HER2+ 10 and hormone receptor positive consists of 3+9+35=47 counts (16.2%), or (1+6+25)/212=15.1% if only Cyprus-born samples are used. (B) ER and PR subtype distribution in BCFR (data taken from [17]. (C) HER2 subtype distribution in BCFR (data taken from [17]. (D) Fisher test p-value of subtype distribution difference between BCFR (or BCAC for the last two rows) and Cyprus cohort (all and Cyprus-born only).

The Fisher’s test for these subtype groupings have been carried out: ER, PR, ER+/PR+ vs ER-/PR-, four ER/PR groups, HER2, triple-negative vs rest, and the p-values are listed in Table 3(D). The lack of significant difference in subtype distribution between North Cyprus cohort and BCFR, indicating certain similarity, is in strong contrast with the dissimilarity of many demographic and reproductive factors as shown in Table 1.

The largest difference is probably in proportion of PR subtypes (higher PR+ proportion in North Cyprus cohort than in BCFR), which may also cause a relatively larger difference for the four ER/PR groups. The highly significant correlation between ER and PR may make PR measurement redundant. In fact, it is argued that added value of PR is questionable [18]. More specifically, ER-/PR+ subtype is rare and may not be reproducible (i.e., can be reclassified to another subtype by another method) [18].

The distribution of breast cancer subtype, defined as the combination of HER2 and ER-PR- (see, e.g. Table 3(C)), is also striking similar between our cohort, the Breast Cancer Association Consortium (BCAC) in UK [19], which consists mostly Europeans/Caucasians. The subtype information is obtained from [20], and Fisher’s p-value for testing triple-negative-only proportion in the two cohorts is not significant (Table 3(D)). Testing four subtypes’ proportions (triple-negative, HER2+only, ER+ and/or PR+ and HER2+, ER+ and/or PR+ and HER2-) is also not significant.

### Predictive factors for breast cancer subtypes

The comparison of factor values between ER+ or PR+ and ER- or PR-samples can also be cast into a regression of ER/PR (dependent variable) over individual factors (independent variables). Table 4 shows all results which are significant at 0.1 level from regressing ER, or PR, or HER2, over either single factor by univariate logistic regression, or all factors by a multiple logistic regression; and for either n=321 samples and for Cyprus-only n=233 samples.

**Table 4.**
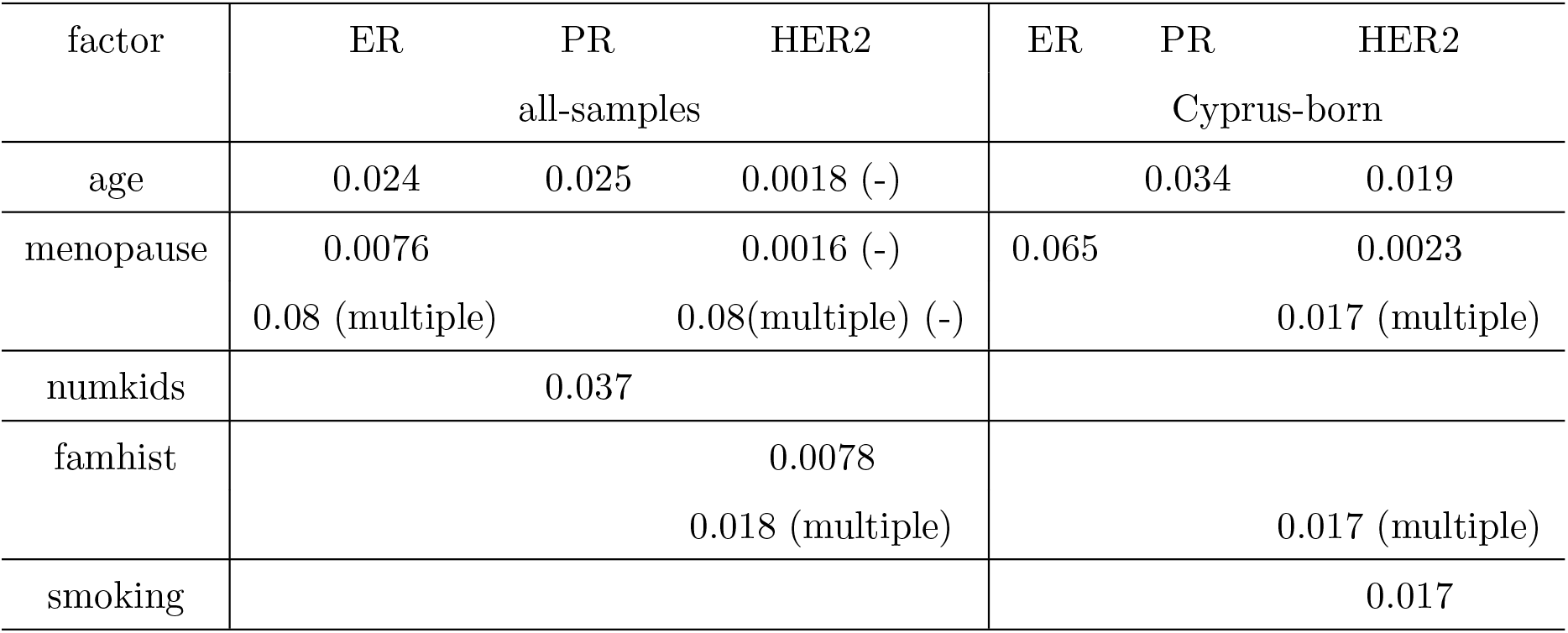
Significant factors in univariate or multiple logistic regression (and the corresponding p-values) Factors that are significantly (at level 0.05) correlated with breast cancer subtypes according to either single-variable or multiple logistic regressions, either by using all samples and by using Cyprus-born only samples. The values are single-variable logistic regression p-values (or multiple variate if marked). “famhist” refers to the presence of any cancer (not necessarily breast cancer) in any first relative.

Table 4 shows age being positively correlated with ER+ and PR+, but negatively correlated with HER2+. These results are similar to what is shown in Table 1 as well as Fig.2 that ER+PR+ patients are older. Due to the positive correlation between ER and PR, ER+ patients and HER2-patients are older.

Table 4 also shows that post-menopause is positively correlated with ER+ but negatively correlated with HER2+. It can be stated that menopause plays a protective role as post-menopause patients are more likely to be in the more curable ER+ subtype and less likely to have the worse prognosis ER-type [21]. The positive correlation between menopause status and age is self-explanatory, and the association between menopause status and ER or HER2 is also easily explained by the age. The last three minor conclusions are: HER2+ patients are more likely to have a first relative with cancer, PR+ patients tend to have more children, and HER2+ patients tend to be smokers.

Between univariate and multiple regression, we also applied three closely related regularized regressions, LASSO (least absolute shrinkage and selection operator), elastic net, and ridge [22], to study the situation with a few explanatory variables. Regularized regression accomplished the task of variable selection (e.g., [23, 24]) by imposing constraint on the sum of absolute value of all fitting coefficients, effectively setting many coefficients to be zero, thus removing the contribution from these variables. Fig.3 shows how the coefficient of each explanatory variable increases, from left to right, when the number of non-zero-coefficient variables increases, for (rows) the dependent variables of ER, ER/PR, HER2, and for (columns) LASSO, elastic net, ridge.

**Figure 3:**
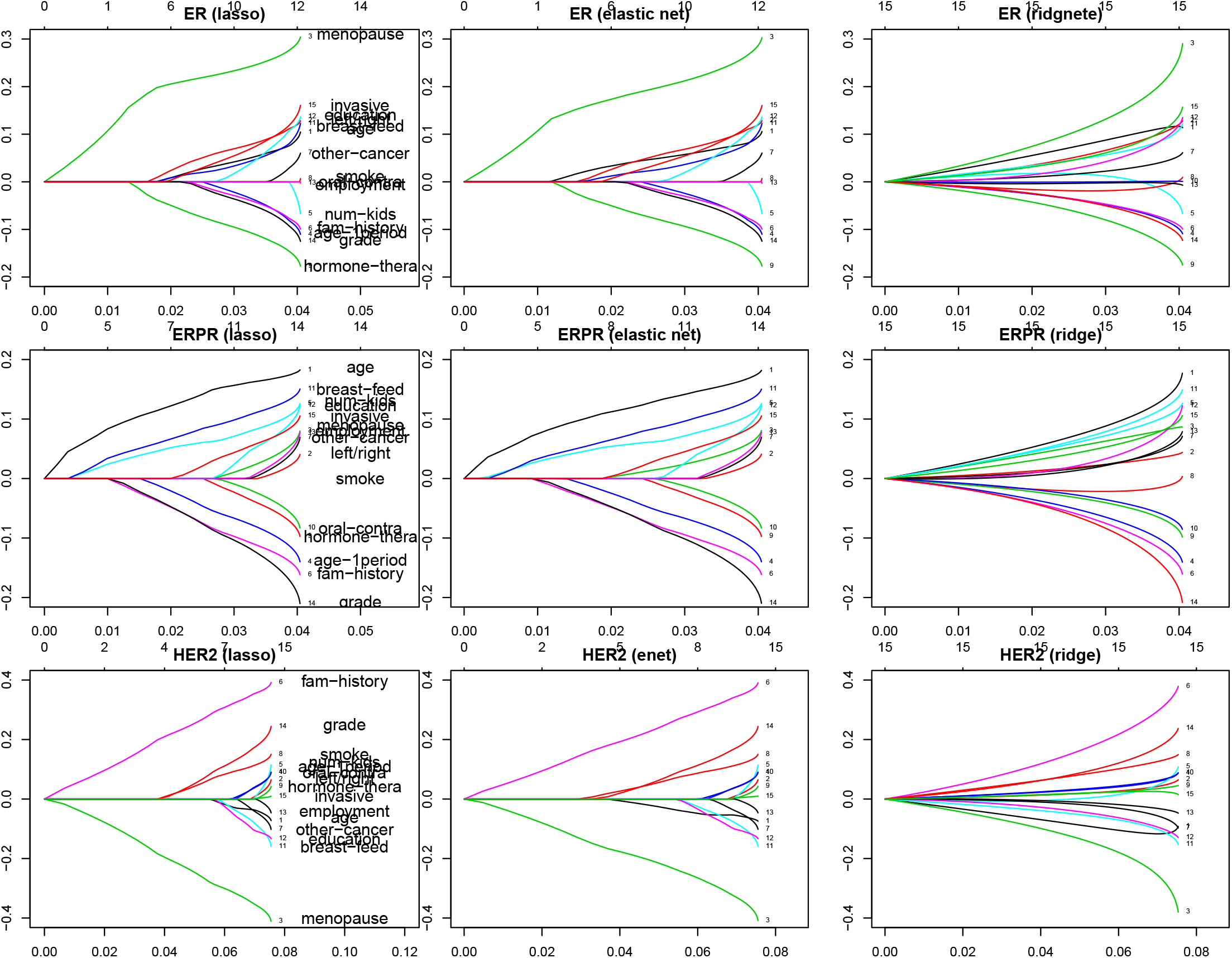
Regularized regression on ER, ER/PR and HER2. Variable tracing/selecting plot of LASSO, elastic net, ridge logistic regressions (columns 1-3) for ER, ER/PR, and HER2 (rows 1-3). Each line is a factor, and positive *x* direction represents a more relaxed constraint, allowing more variables. The *y* axis is the coefficient of a factor/variable: positive (negative) coefficient means a positive (negative) correlation between the factor and the subtype status (ER+, ER+PR+, HER+ are 1’s, ER-, ER-PR-, HER2-are 0’s). The *x* axis is deviance explained by the (regularized) logistic regression.

First of all, we see that LASSO, elastic net, and ridge regression show similar trend with the same breast cancer subtype. Therefore, we may only focus on LASSO plot, which are the first column in Fig.3. Secondly, in order to see how n=321 dataset may differ from the n=233 Turkish-Cypriots only dataset, we draw the LASSO results side by side in Fig.4.

**Figure 4:**
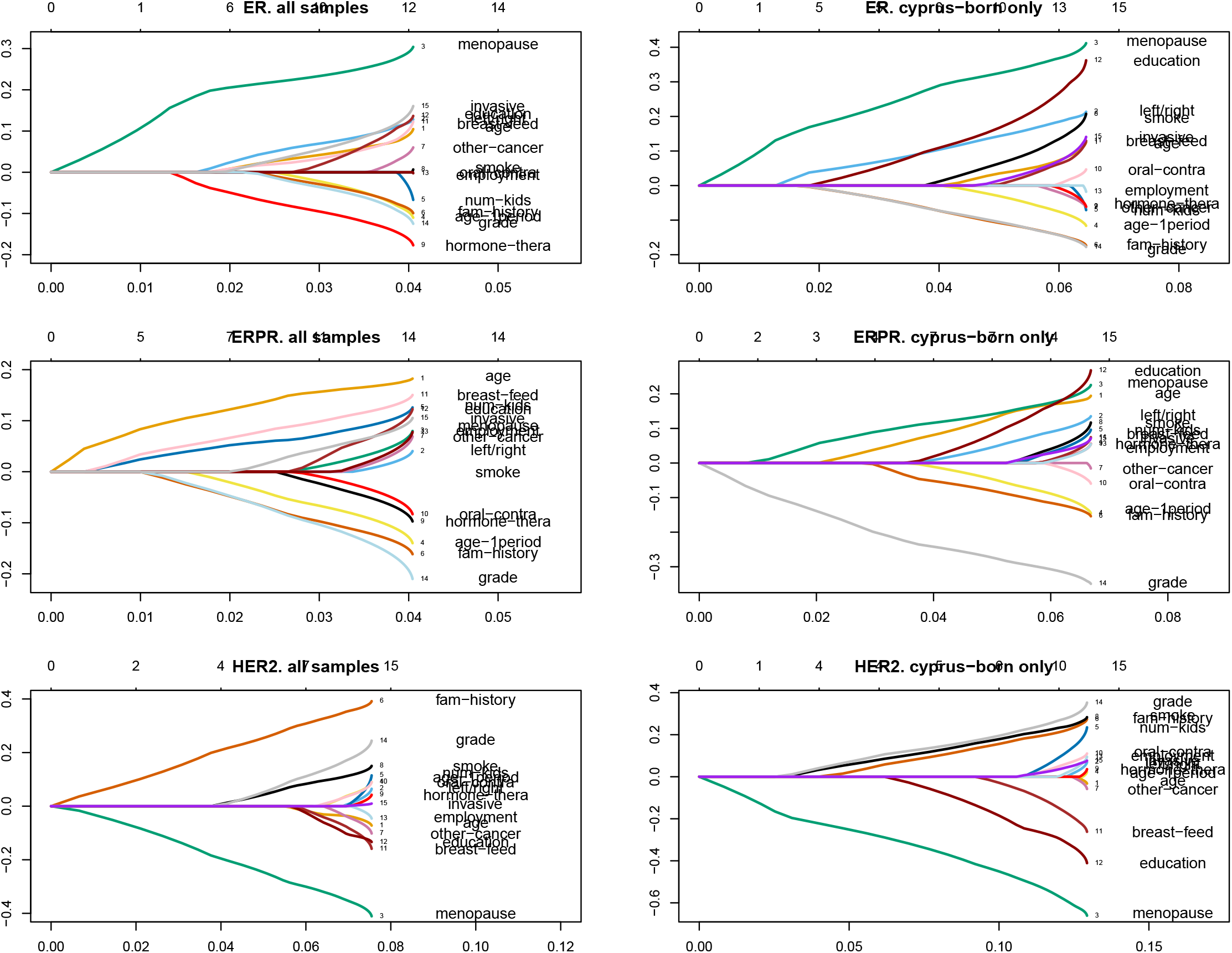
Comparison of LASSO regression between n=321 and n=233 sets. LASSO regression results for all n=321 patients (left) and for n=233 Turkish Cypriot patients (right), for ER, ER/PR, HER2 (rows 1-3). See Fig.3 for more explanation of the x and y axes.

For ER subtype, the dominant contribution from menopause status is not only consistently shown for both n=321 and n=233 datasets, but also consistent with Table 4. The hormone therapy is a promising signal for n=321 dataset but not so for n=233 one. For ER/PR, the cancer stage is a consistent signal which is negatively correlated with the ER+/PR+ status. This signal is also highlighted in BCFR paper [17]. Breast-feeding is a promising signal for n=321 dataset, but seem to be less important for n=233 dataset. More discussion on the benefit of breast-feeding in reducing the probability in acquiring poor prognostic breast cancers, such as triple negative subtypes, can be found in [25, 26]. For HER2 subtype, similar to univariate and multiple regression (Table 4), menopause is a dominant factor with negative correlation. Interestingly, there is a signal for n=321 dataset from first-degree-relative cancer history, but that signal becomes weaker for the n=233 dataset.

## Discussion

Without control samples, we carried out a case-only analysis of potential predictive factors of different subtypes of breast cancer. Case-only design has been implemented in breast cancer studies before, and it is “an important initial step in understanding the extent of etiologic heterogeneity between tumor subtypes” [10, 11]. Since different subtypes of breast cancer have different prognostics, it is important to assess their distribution.

One of the striking results we obtained is that our North Cyprus has very similar ER+, ER+/PR+, and HER2+ as BCFR, even though our cohort is much older, more post-menopause, less educated, less hormone therapy use, more breastfeeding, etc. If older/post-menopause patients tend to have ER+, and our older cohort should have higher proportion of ER+ subtype than BCFR. Although it is indeed the case (74.5% ER+ in n=321 Cyprus dataset and 71.9% ER+ in BCFR samples), the difference on underlying factors (age or menopause status) is very significant between the two cohorts, but not significant for ER distributions.

This topic can be discussed in a general term: can correlation at one level be translated to correlation at another level? In our case, we examine the potential similarity/dissimilarity of distribution of a factor in two data sets (low-level), and wonder whether it can be translated to the similarity/dissimilarity of distribution of a subtype affected by these predictive factor (high-level) in those two datasets. In our previous investigation of a very different issue, linkage/association analysis of multiple correlated phenotypes in a lipid panel, we have observed that correlation at the high-level (phenotype) does not necessarily translate to a correlation at the low-level (linkage-disequilibrium or co-localization between genetic variants) [27].

It could also be that the causal link between the two levels are not strong enough to transfer correlation from one level to another. In our LASSO analysis (Fig.3 and 4), it can be seen that the fraction of deviation explained (range of x axis) of ER, ER/PR, HER2 is at most a few percent, even using all factors. Random forest run on the same data also show that the classification rate on ER, or ER/PR, or HER2 status is not high: on average barely over 50% (results not shown). It highlights the fact that many true predictive factors of breast cancer subtypes are not yet included in our data (e.g., body mass index (BMI)), and also the known genetic causes of breast cancer (e.g., BRCA1 and BRCA2) is not part of the analysis.

In a recent systematic meta analysis of African breast cancer subtypes [28], it is found that proportion of ER+ and PR+ samples fluctuate greatly from study to study. There are also data showing the triple-negative subtype rate is much higher in African women than European/Caucasian [29, 30]. To double check whether breast cancer subtype distribution in our North Cyprus cohort is still the same with another study, we picked a published summary statistics from a southeastern Turkish cohort [31]. The ER+, ER+/PR+, HER2+ proportions in the Turkish cohort are 73.5%, 81.8%, 30.4%, compared to our 74.5%, 75.9% and 24.1%, leading to Fisher test p-values of 0.8, 0.086, and 0.076 (number of sample in Turkish cohort with the subtype information are 438, 437, 434). These differences are within ranges, and are not significant.

It could be of great interest to compare our breast cancer subtypes statistics with a Greek Cypriots cohort. We found two surveys on breast cancer in Greek Cypriots, one with 1100 patients conducted from 1999 to 2005 [32], and another more than 4000 patients conducted from year 2005 to 2013 [33]. Unfortunately, there is no hormone receptor subtype information for the Greek Cypriots cohort in both time periods. However, we could compare the distribution of other factors when the comparable data is available. We found that for 1995-2005 period, number of children distribution is almost identical between the Greek and Turkish Cypriot cohorts; age, smoking status are not significantly different; Greek Cypriot patients have higher education level (p-value=1E-8), less family breast cancer history (p-value=5E-7), more early (less or equal 11 years old) menarche (p-value=0.04), less breastfeeding (p-value= 0.02), less use of oral contraceptive (p-value= 0.004). For 2005-2013 data, smoking status difference is not significant, and there are more invasive patients in Greek Cypriots than Turkish Cypriots (p-value=1E-14).

Our regularized regression (LASSO, elastic net and ridge) (Fig.3, 4) reveal potential predictive factors to breast cancer subtypes. However, these weak signals can only be considered to be “trend” which is not yet proven by statistical test as those in Table 4. In [34], benign disease proliferation risk is higher in ER+ subtype patients than in ER-group. This can be compared our positive contribution from other cancer (including metastasis) and ER+, or ER+/PR+ subtype (Fig.4). In [17], not breastfeeding is associated with ER-/PR-subtype, which can be compared with our result that breastfeeding is positively correlated with the ER+/PR+ subtype. In [21], ER-cancer rate stop to increase at certain age, whereas ER+ rate continue to increase. This can be compared to our result that post-menopause is is positively correlated with the ER+ subtype (Fig.4). In [35], significant difference of age, menopause status, past use of hormone therapy is observed in four ER/PR group. In [36], early age at menarche (≤ 12 years) is less common in PR-group than PR+ group, and it is also true in our data comparing ER-/PR- and ER+/PR+ groups. To summarize, many of our observed predictive factors for breast cancer subtypes are consistent with the literature. The positive correlation between cancer family history and HER2+ subtype (Fig.4) remains intriguing.

In conclusion, we use a unique cohort of breast cancer in a under-studied population to survey the breast cancer subtypes and related factors. We use a simplified analysis framework: keeping breast cancer subtypes at one level, and all factors at another level. Distribution of many factors are extremely different from that of another large breast cancer registry, while the subtype distribution is similar. This indirectly shows that we have not exhaustively measured all predictive factors of breast cancer subtypes. The relationship between the two levels is investigated by regression, with one variable, all variable, or subset of variables. These regression analyses show post-menopause and/or older breast cancer patients are more likely to have the ER+ subtype and HER2-subtype. We have also observed several other trends which should be validated in a larger cohort.

## Data Availability

individual person's data is not available. summary data is available upon request.

## Acknowledgements

This study was supported through the Fulbright Visiting Research Scholarship Grant by the US Department of State. AU would like to thank her advisor Prof. Mary Terry Beth and her colleagues at the BCFR at the Department of Epidemiology at Columbia University in New York for their help and support. AU would also like to thank to the Ministry of Health at the Turkish Republic of Northern Cyprus for access to the breast cancer archives at the Burhan Nalbantoglu State Hospital in Nicosia, and Dr. Nilay Acar, Dr. Mehmet Ali Alpdoğan, Dr. Fuat Ağlarcan, and Dr.Whitney A. Onuorah from the Faculty of Medicine, Eastern Mediterranean University for their help and guidance for the data collection. WT would like to thank the support from Robert S Boas Center for Genomics and Human Genetics.

## References

[1] J Ferlay, M Colombet, I Soerjomataram, T Dyba, G Randi, M Bettio, A Gavin, O Visser, F Bray (2018), Cancer incidence and mortality patterns in Europe: Estimates for 40 countries and 25 major cancers in 2018, Euro. J. Cancer, 103:356–287.

[2] K McPherson, CM Steel, JM Dixon (2000), Breast cancerepidemiology, risk factors, and genetics, BMJ, 321:624–628.

[3] S Möller, LA Mucci, JR Harris, T Scheike, K Holst, U Halekoh, HO Adami, K Czene, K Christensen, NV Holm, E Pukkala, A Skytthe, J Kaprio, JB Hjelmborg (2016), The heritability of breast cancer among women in the Nordic twin study of cancer, Cancer Epidemiol. Biomarkers & Prev., 25:145–150.

[4] N Mavaddat, K Michailidou, J Dennis, M Lush, L Fachal, A Lee, JP Tyrer, TH Chen, Q Wang, MK Bolla, et al. (2019), Polygenic risk scores for prediction of breast cancer and breast cancer subtypes, Am. J. Hum. Genet., 104:21–34.

[5] Y Shieh, L Fejerman, PC Lott, K Marker, SD Sawyer, D Hu, S Huntsman, J Torres, M Echeverry, ME Bohorquez, et al. (2019), A polygenic risk score for breast cancer in U.S. Latinas and Latin-American women, bioRxiv preprint 598730. DOI: 10.1101/598730

[6] A Lee, N Mavaddat, AN Wilcox, AP Cunningham, T Carver, S Hartley, CB de Villiers, A Izquierdo, J Simard, MK Schmidt, FM Walter, N Chatterjee, M Garcia-Closas, M Tischkowitz, P Pharoah, DF Easton, AC Antoniou (2019), BOADICEA: a comprehensive breast cancer risk prediction model incorporating genetic and nongenetic risk factors, Genet. in Med., in press.

[7] ER Fisher, CK Redmond, H Liu, H Rockette, B Fisher (1980), Correlation of estrogen receptor and pathologic characteristics of invasive breast cancer, Cancer, 45:349–353.

[8] BD Lehmann, JA Bauer, X Chen, ME Sanders, AB Chakravarthy, Y Shyr, JA Pietenpol (2011), Identification of human triple-negative breast cancer subtypes and preclinical models for selection of targeted therapies, J. Clin. Invest., 121:2750–2767.

[9] V özmen, T özmen, V Doğru (2019), Breast cancer in Turkey; an analysis of 20,000 patients with breast cancer, Euro. J. Breast Health, 15:141–146.

[10] M. Martínez, GI Cruz, AM Brewster, ML Bondy, PA Thompson (2010), What can we learn about disease etiology from case-case analyses? lessons from breast cancer, Cancer Epid. Biomarker & Prev., 19:2710–2714.

[11] CM Redondo, M Gago-Domínguez, SM Ponte, ME Castelo, X Jiang, AA Garca, M. Fernández, MA Tomé\, M Fraga, F Gude, ME Martnez, VM Garzn, Carracedo, JE Castelao (2012), Breast feeding, parity and breast cancer subtypes in a Spanish cohort, PLoS ONE, 7:e40543.

[12] J Friedman, T Hastie, R Tibshirani (2010), Regularization paths for generalized linear models via coordinate descent, J. Stat. Software, 33:1–22.

[13] LJ Van der Maaten and GE Hinton (2008), Visualizing data using t-SNE, J. Machine Learning Res., 9:2575–2605.

[14] W Li, JE Cerise, Y Yang, H Han (2017), Application of t-SNE to human genetic data, J. Bioinfo. and Comp. Biol., 15:1750017.

[15] W Li, J Freudenberg, J Freudenberg (2019), Alignment-free approaches for predicting novel Nuclear Mitochondrial Segments (NUMTs) in the human genome, Gene, 691:141–152.

[16] HJ Switf, D Abrams, L Drury, RA Lamont (2018), Categorization by age, in Encyclopedia of Evolutionary Psychological Science, eds. T Shackelford, V Weekes-Shackelford (Springer). doi: 10.1007/978-3-319-16999-62431-1

[17] ME Work, EM John, IL Andrulis, JA Knight, Y Liao, AM Mulligan, MC Southey, GG Giles, GS Dite, C Apicella, H Hibshoosh, JL Hopper, MB Terry (2014), Reproductive risk factors and oestrogen/progesterone receptor-negative breast cancer in the Breast Cancer Family Registry, Brit. J. Cancer, 110:1367–1377.

[18] MM Hefti, R Hu, NW Knoblauch, LC Collins, B Haibe-Kains, RM Tamimi, AH Beck (2013), Estrogen receptor negative/progesterone receptor positive breast cancer is not a reproducible subtype, Breast Cancer Res., 15:R68.

[19] Breast Cancer Association Consortium (2006), Commonly studied single-nucleotide polymorphisms and breast cancer: results from the Breast Cancer Association Consortium, N. Natl. Cancer Inst., 98:1382–1396.

[20] O Brouckaert, A Rudolph, A Laenen, R Keeman, MK Bolla, Q Wang, A Soubry, H Wildiers, IL Andrulis, V Arndt, et al. (2017), Reproductive profiles and risk of breast cancer subtypes: a multi-center case-only study, Breast Cancer Res., 19:119.

[21] RE Tarone and KC Chu (2002), The greater impact of meonopause on ER-than ER+ breast cancer incidence: a possible explanation, Cacner Causes & Control, 13:7–14.

[22] T Hastie, R Tibshirani, J Friedman (2009), The Elements of Statistical Learning, 2nd ed (Springer).

[23] RS Halinski and LS Feldt (1970), The selection of variables in multiple regression analysis, J. Edu. Measurement, 7:151–157.

[24] W Li and Y Yang (2002), How many genes are needed for a discriminant microarray data analysis, in Methods of Microarray Data Analysis, eds. SM Lin, KF Johnson, (Kluwer Acvademic Publishers), pp.137–149.

[25] F Islami, Y Liu, A Jemal, J Zhou, E Weiderpass, G Colditz, P Boffetta, M Weiss (2015), Breastfeeding and breast cancer risk by receptor status – a systematic review and meta-analysis, Ann. Oncol., 26:2398–2407.

[26] RT Fortner, J Sisti, B Chai, LC Collins, B Rosner, SE Hankinson, RM Tamimi, AH Eliassen (2019), Parity, breastfeeding, and breast cancer risk by hormone receptor status and molecular phenotype: results from the Nurses Health Studies, Breast Cancer Res., 21:40.

[27] A Ulgen, Z Han, W Li (2003), Correlation between quantitative traits and correlation between corresponding LOD scores: detection of pleiotropic effects, BMC Genet., 4:S60.

[28] A Eng, V McCormack, I Dos-Santos-Silva (2014), Receptor-defined subtypes of breast cancer in indigenous populations in Africa: a systematic review and meta-analysis, PLoS Med., 11:e1001720.

[29] D Huo, F Ikpatt, A Khramtsov, J-M Dangou, R Nanda, J Dignam, B Zhang, T Grushko, C Zhang, O Oluwasola, D Malaka, S Malami, A Odetunde, AO Adeoye, F Iyare, A Falusi, CM Perou, OI Olopade (2009), Population differences in breast cancer: survey in indigenous African women reveals over-representation of triple-negative breast cancer, J. Clin. Oncol., 27:4515–4521.

[30] Y Zheng, T Walsh, S Gulsuner, S Casadei, MK Lee, TO Ogundiran, A Ademola, AG Falusi, CA Adebamowo, AO Oluwasola, A Adeoye, A Odetunde, CP. Babalola, OA Ojengbede, S Odedina, I Anetor, S Wang, D Huo, TF Yoshimatsu, J Zhang, GES Felix, M-C King, OI Olopade (2018), Inherited breast cancer in Nigerian women, J. Clin. Oncol., 36:2820–2825.

[31] A Kuzhan, M Adli, H Eryigit Alkis, D Caglayan (2013), Hormone receptor and HER2 status in patients with breast cancer by races in southeastern Turkey, J. Balkan Union Oncol., 18:619–622.

[32] A Hadjisavvas, MA Loizidou, N Middleton, T Michael, R Papachristoforou, E Kakouri, M Daniel, P Papadopoulos, S Malas, Y Marcou, K Kyriacou (2010), An investigation of breast cancer risk factors in Cyprus: a case control study, BMC Cancer, 10:447.

[33] P Pilavaki, G Giallouros, AI Yiallourou, K Pantavou, Y Marcou, A Demetriou, V Scoutellas, GK Nikolopoulos (2018), Epidemiology of breast cancer in Cyprus: Data on newly diagnosed cases and survival rates, Data Brief, 19:353–369.

[34] K Kerlikowske, CC Gard, JA Tice, E Ziv, SR Cummings, DL Miglioretti, Breast Cancer Surveillance Consortium (2016), Risk factors that increase risk of estrogen receptor-positive and -negative breast cancer, J. Natl. Cancer Inst., 109:djw276.

[35] GA Colditz, BA Rosner, WY Chen, MD Holmes, SE Hankinson (2004), Risk factors for breast cancer according to estrogen and progesterone receptor status, J. Natl. Cancer Inst., 96:218–228.

[36] XR Yang, J Chang-Claude, EL Goode, FJ Couch, H Nevanlinna, RL Milne, M Gaudet, MK Schmidt, A Broeks, A Cox, et al. (2010), Associations of breast cancer risk factors with tumor subtypes: a pooled analysis from the Breast Cancer Association Consortium studies, J. Natl. Cancer Inst.,103:250–263.

[37] D Colquhoun (2014), An investigation of the false discovery rate and the misinterpretation of p-values, Royal Soc. Open Sci., 1:140216.

